# Delirium and Neuropsychological Outcomes in Critically Ill Patients with COVID-19: an Institutional Case Series

**DOI:** 10.1101/2020.11.03.20225466

**Authors:** Jacqueline Ragheb, Amy McKinney, Mackenzie Zierau, Joseph Brooks, Maria Hill-Caruthers, Mina Iskander, Yusuf Ahmed, Remy Lobo, Graciela Mentz, Phillip E. Vlisides

## Abstract

**Objective:** To characterize the clinical course of delirium for COVID-19 patients in the intensive care unit, including post-discharge cognitive outcomes.

**Patients and Methods:** A retrospective chart review was conducted for patients diagnosed with COVID-19 (n=148) admitted to an intensive care unit at Michigan Medicine between March 1, 2020 and May 31, 2020. A validated chart review method was used to identify presence of delirium, and various measures (e.g., Family Confusion Assessment Method, Short Blessed Test, Patient-Health Questionnaire-9) were used to determine neuropsychological outcomes between 1-2 months after hospital discharge.

**Results:** Delirium was identified in 108/148 (73%) patients in the study cohort, with median (interquartile range) duration lasting 10 (4 – 17) days. In the delirium cohort, 50% (54/108) of patients were African American, and delirious patients were more likely to be female (76/108, 70%) (absolute standardized differences >.30). Sedation regimens, inflammation, deviation from delirium prevention protocols, and hypoxic-ischemic injury were likely contributing factors, and the most common disposition for delirious patients was a skilled care facility (41/108, 38%). Among patients who were delirious during hospitalization, 4/17 (24%) later screened positive for delirium at home based on caretaker assessment, 5/22 (23%) demonstrated signs of questionable cognitive impairment or cognitive impairment consistent with dementia, and 3/25 (12%) screened positive for depression within two months after discharge.

**Conclusion:** Patients with COVID-19 commonly experience a prolonged course of delirium in the intensive care unit, likely with multiple contributing factors. Furthermore, neuropsychological impairment may persist after discharge.

## Introduction

The outbreak of Severe Acute Respiratory Syndrome Coronavirus 2 (SARS-CoV-2), the virus that causes Coronavirus Disease (COVID-19), emerged as a public health threat in December 2019 and was declared a pandemic by World Health Organization in March 2020. Major neurological complications, such as encephalopathy, delirium, strokes, seizures, and ataxia, have all been observed.^1-5^ Delirium appears to be a common complication, with previous investigations demonstrating an incidence of approximately 65-80% in the intensive care unit (ICU).^1, 4^ Delirium may occur due to direct coronavirus invasion of the central nervous system, ^6^ and systemic inflammatory responses may further exacerbate neurocognitive impairment. In the ICU, synergistic factors such as sedation regimen, social isolation, and deviation from standard care protocols may further increase risk. Delirium is also associated with prolonged hospitalization, long-term cognitive and functional impairment, and increased mortality.^7-9^ As such, there is a critical need to improve understanding of this syndrome in patients with COVID-19.

While a high incidence of delirium has been reported in COVID-19 patients, fundamental questions persist. The clinical course of delirium, including average duration and post-discharge cognitive trajectory, remains unknown. Pathophysiologic drivers of delirium are incompletely understood, and the extent to which standard prevention protocols are implemented is unclear. Such detailed understanding will contribute to delirium phenotyping of COVID-19 patients and provide insight into the clinical and neurocognitive burden associated with COVID-19. In this context, the objective of this study was to determine granular details associated with delirium in ICU patients with COVID-19. Specifically, the clinical course of delirium, presence of exacerbating factors, nature of prevention strategy implementation, and post-discharge cognitive outcomes were all characterized.

## Methods

### Study design and overview

This was a single-center case series from Michigan Medicine. Detailed chart review data were collected from critically ill patients with COVID-19 (3/1/2020 – 5/31/2020), and post-discharge telephone surveys were conducted to test neuropsychological function after discharge. All study operations were conducted at Michigan Medicine, Ann Arbor MI USA, and approval was obtained from the University of Michigan Medical School Institutional Review Board (HUM00182646). A Health Insurance Portability and Accountability Act waiver was granted in order to retrospectively review patient medical records, and informed consent was not required for retrospective chart review. Patients who agreed to complete telephone surveys after discharge were consented over the telephone prior to survey administration using a comprehensive consent document. A waiver of documentation of consent was approved in conjunction with Institutional Review Board approval and as required by U.S. Department of Health and Human Services regulations and policy. Lastly, the CAse REport guidelines checklist is included in the supplemental online material (Supplemental Table 1). These guidelines provide reporting standards for case reports of one or more patients.^10^

**Table 1.** Baseline Characteristics

|  | **All Patients**<br>**(n=148)** | **Delirium**<br>**(n=108)** | **No Delirium**<br>**(n=40)** | **Absolute**<br>**Standardized**<br>**Difference** |
| --- | --- | --- | --- | --- |
| **Age (IQR)** | 59 (49 – 71) | 58 (47 – 71) | 62 (54 – 71) | .26 |
| **Male sex, n (%)** | 98 (66) | 32 (30) | 18 (45) | .32 |
| **Race, n (%)** |  |  |  | .32 |
| Caucasian | 66 (45) | 47 (44) | 19 (48) |  |
| African-American | 70 (47) | 54 (50) | 16 (40) |  |
| Other | 1 (0.7) | 1 (0.9) | 0 (0) |  |
| Not reported | 11 (7.4) | 6 (5.6) | 5 (13) |  |
| **Ethnicity, n (%)** |  |  |  | .31 |
| Non-Hispanic | 137 (93) | 100 (93) | 38 (93) |  |
| Hispanic | 5 (3.4) | 5 (4.7) | 0 (0) |  |
| Unknown/not<br>reported | 6 (4.1) | 3 (2.8) | 3 (7.5) |  |
| **Weight, kg (IQR)** | 103 (83 – 127) | 105 (87 – 127) | 93 (79 – 113) | .36 |
| **BMI (IQR)** | 34 (28 – 40) | 34 (29 – 41) | 31 (28 – 39) | .16 |
| **Comorbidities,**<br>**n (%)** |  |  |  |  |
| Asthma | 24 (16) | 17 (16) | 7 (18) | .05 |
| Atrial fibrillation | 22 (15) | 14 (13) | 8 (20) | .19 |
| Cancer | 25 (17) | 20 (19) | 5 (13) | .17 |
| Chronic kidney<br>disease | 40 (27) | 30 (28) | 10 (25) | .06 |
| Congestive heart<br>failure | 19 (13) | 13 (12) | 6 (15) | .09 |
| COPD | 14 (9.5) | 8 (7.4) | 6 (15) | .24 |
| Coronary artery disease | 27 (18) | 18 (17) | 9 (23) | .15 |
| Depression | 17 (11) | 11 (10) | 6 (15) | .15 |
| Diabetes mellitus | 75 (51) | 58 (54) | 18 (43) | .23 |
| Hypertension | 102 (69) | 74 (66) | 29 (70) | .03 |
| Obstructive sleep apnea | 31 (21) | 22 (20) | 9 (23) | .05 |
| Seizures | 8 (5.4) | 5 (4.6) | 3 (7.5) | .12 |
| Stroke | 9 (6.1) | 5 (4.6) | 4 (10) | .21 |
| Substance abuse | 9 (6.1) | 6 (5.6) | 3 (7.5) | .21 |
| TIA | 5 (3.4) | 3 (2.8) | 2 (5) | .12 |

Median (interquartile range, IQR) data presented. *Kg* kilograms, *BMI* body mass index, *COPD* chronic obstructive pulmonary disease, *TIA* transient ischemic attack.

### Eligibility criteria

All patients with a COVID-19 diagnosis admitted to a Michigan Medicine ICU between 03/01/2020 – 05/31/2020 were eligible for study inclusion. ICU patients admitted during this time, without a diagnosis of COVID-19, were not eligible for study inclusion.

### Outcomes

The primary outcome was delirium presence (yes/no, %) at any point during admission. Delirium was evaluated via chart review method (described below). Several secondary outcomes were also collected in relation to delirium and overall clinical trajectory. These outcomes included the following: duration of delirium (days), antipsychotic administration, length of hospital stay, length of ICU stay, number of days requiring ventilator support, inflammatory laboratory values (white blood cell count, c-reactive protein, ferritin, lactate dehydrogenase, d-dimer, and interleukin-6), new psychiatry consults, new antidepressant use, and final disposition (e.g., home, long-term care facility, death). Delirium prevention strategies, based on the ABCDEF ICU liberation bundle,^11, 12^ were also recorded. These included the following: structured mobility exercises, placing familiar objects from home at the bedside, promoting use of visual and hearing aids, and spontaneous awakening/breathing trials. The total number of times a prevention strategy was charted was recorded for each patient, and this number was divided by the *expected* number of times that intervention should have occurred based on length of ICU stay and protocolized schedule. This provided the estimated compliance rate for each intervention. Neuroimaging data were also collected and reviewed.

Lastly, a telephone survey was conducted between 30-60 days post-discharge to determine whether subjective or objective signs of cognitive impairment were present. During telephone interviews, the following tests were conducted: the Patient-Reported Outcomes Measurement Information System (PROMIS)^13^ Cognitive Function Abilities 4a, Short Blessed Test (score 0-4 = normal cognition, score 5-9 = questionable impairment, score ≥10 = impairment consistent with dementia),^14^ Family Confusion Assessment Method (FAM-CAM) for delirium,^15^ and the Patient Health Questionnaire-9 (scores ≥10 were considered positive screens for depression).^16, 17^

### Data collection

Screening for eligible patients was first performed for via DataDirect, a software tool from the University of Michigan Office of Research that enables research teams to retrospectively search for patient cohorts. Charts that screened positive were then manually reviewed by study team members to confirm study eligibility.

Charts were then reviewed in further detail for outcome abstraction. Delirium was assessed via validated, publicly available chart review method.^18^ Briefly, any instance of an acute confusional state was recorded in the instrument and counted as an episode of delirium. The methodology is drawn from the Confusion Assessment Method,^19^ which assesses for acute changes in cognition, fluctuating course, inattention, altered levels of consciousness, and disorganized thinking. This was the core set of delirium symptoms in this cohort, and hyperactive states (e.g., agitation) were reported as well. The source of information was recorded, along with the date and time. The total number of days with acute confusion was also included in the instrument, along with any evidence of reversibility or improvement of the confusion state. Other clinical outcomes, along with laboratory values, were collected directly from the charts. Neuroimaging studies were manually reviewed by a board certified radiologist with a Certificate of Added Qualification in neuroradiology (R.L.).

### Statistical Analysis

Analyses were performed using IBM SPSS version 27 (Armonk, NY USA) and SAS version 9.4 (SAS Institute, Cary, NC USA). Exploratory data analysis techniques were used to assess the distribution of dependent measures for determining the appropriate analytical strategy. The Shapiro-Wilk test was used to assess the distribution of continuous outcomes, and Independent t-tests or Mann-Whitney U tests were used as appropriate. Mean (standard deviation) or median (interquartile range) was reported for parametric and non-parametric data, respectively. For binary outcomes and proportions, The Chi-Square Test or Fisher’s Exact Test were used, as appropriate. Absolute standardized differences were calculated for determining differences in baseline characteristics between groups, with differences >.20 considered to be imbalanced. The threshold for significance was set to p<0.05 across all tests otherwise. For post-discharge cognitive outcomes, descriptive statistics were reported.

## Results

Baseline characteristics are presented in Table 1. The majority of patients were African-American and non-Hispanic, and the most common comorbidities were hypertension, Diabetes mellitus, and obesity. Absolute standardized differences between delirium and non-delirium groups were largest (>.30) for sex, race, and weight. The highest proportion of patients in the delirium group were African-American (n=54, 50%), and weight was significantly higher in the delirium group (105 [87 – 127] kg vs. 93 [97 – 113] kg, *P*<.05).

### Delirium and Neuropsychological Outcomes

Delirium incidence was high in the cohort (108/148, 73%), and median (interquartile range) duration was 10 (4 – 17) days (Table 2). Delirium prevention activities occurred relatively infrequently, with estimated unit protocol compliance rates less than 50% for each intervention reported (see Table 2 legend for description of protocol activity schedule). The mobility exercise activity compliance rate (%) was significantly lower in the delirium group (37% [26 – 55]) compared to the non-delirium group (62% [31 – 152]; *P*=.009). Likewise, daily promotion of visual and hearing aids occurred less frequently in the delirium group (27% [13 – 63]) compared to the non-delirium group (77% [14 – 213]; *P*=.005). New antidepressant use was more common for those with delirium (27/108, 25%) compared to those without delirium (3/40, 7.5%; *P*=.01). Similarly, a psychiatry consult was obtained for 21/108 (19%) delirious patients compared to 0/40 (0%) in the non-delirium group (*P*=.003). Lastly, no evidence of reversal or improvement was reported for more than 30% of patients during index hospitalization.

**Table 2.**
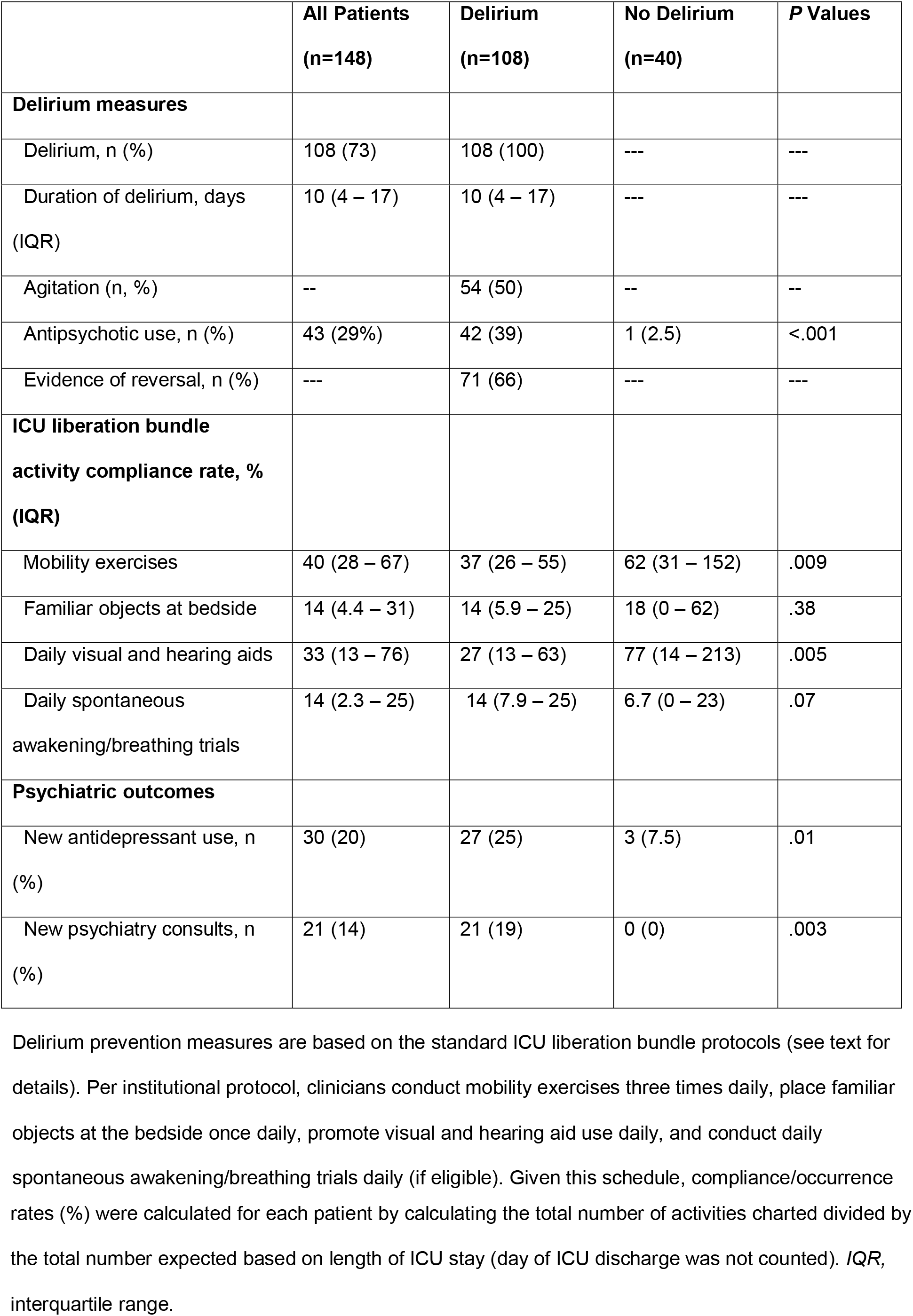
Delirium and Neuropsychological Outcomes

|  | **All Patients**<br>**(n=148)** | **Delirium**<br>**(n=108)** | **No Delirium**<br>**(n=40)** | **P Values** |
| --- | --- | --- | --- | --- |
| **Delirium measures** |  |  |  |  |
| Delirium, n (%) | 108 (73) | 108 (100) | --- | --- |
| Duration of delirium, days<br>(IQR) | 10 (4 – 17) | 10 (4 – 17) | --- | --- |
| Agitation (n, %) | -- | 54 (50) | -- | -- |
| Antipsychotic use, n (%) | 43 (29%) | 42 (39) | 1 (2.5) | <.001 |
| Evidence of reversal, n (%) | --- | 71 (66) | --- | --- |
| **ICU liberation bundle**<br>**activity compliance rate, %**<br>**(IQR)** |  |  |  |  |
| Mobility exercises | 40 (28 – 67) | 37 (26 – 55) | 62 (31 – 152) | .009 |
| Familiar objects at bedside | 14 (4.4 – 31) | 14 (5.9 – 25) | 18 (0 – 62) | .38 |
| Daily visual and hearing aids | 33 (13 – 76) | 27 (13 – 63) | 77 (14 – 213) | .005 |
| Daily spontaneous<br>awakening/breathing trials | 14 (2.3 – 25) | 14 (7.9 – 25) | 6.7 (0 – 23) | .07 |
| **Psychiatric outcomes** |  |  |  |  |
| New antidepressant use, n<br>(%) | 30 (20) | 27 (25) | 3 (7.5) | .01 |
| New psychiatry consults, n<br>(%) | 21 (14) | 21 (19) | 0 (0) | .003 |

Delirium prevention measures are based on the standard ICU liberation bundle protocols (see text for details). Per institutional protocol, clinicians conduct mobility exercises three times daily, place familiar objects at the bedside once daily, promote visual and hearing aid use daily, and conduct daily spontaneous awakening/breathing trials daily (if eligible). Given this schedule, compliance/occurrence
rates (%) were calculated for each patient by calculating the total number of activities charted divided by the total number expected based on length of ICU stay (day of ICU discharge was not counted). *IQR*, interquartile range.

### Hospitalization and Post-Discharge Outcomes

Median length of hospitalization was 25 (13 – 48) days, and median length of ICU stay was 15 (7 – 31) days across the cohort (Table 3). Length of hospitalization, ICU length of stay, and duration of mechanical ventilation were all significantly prolonged in patients experiencing delirium (Table 3). Correspondingly, sedative-hypnotic use was higher in patients with delirium. Delirious patients demonstrated higher white blood cell counts, c-reactive protein levels, and d-dimer levels compared to non-delirious patients. Less than half of patients were ultimately discharged home, and the most common disposition for those with delirium was a skilled care facility (41/108, 38%) after discharge (Table 3).

**Table 3.**
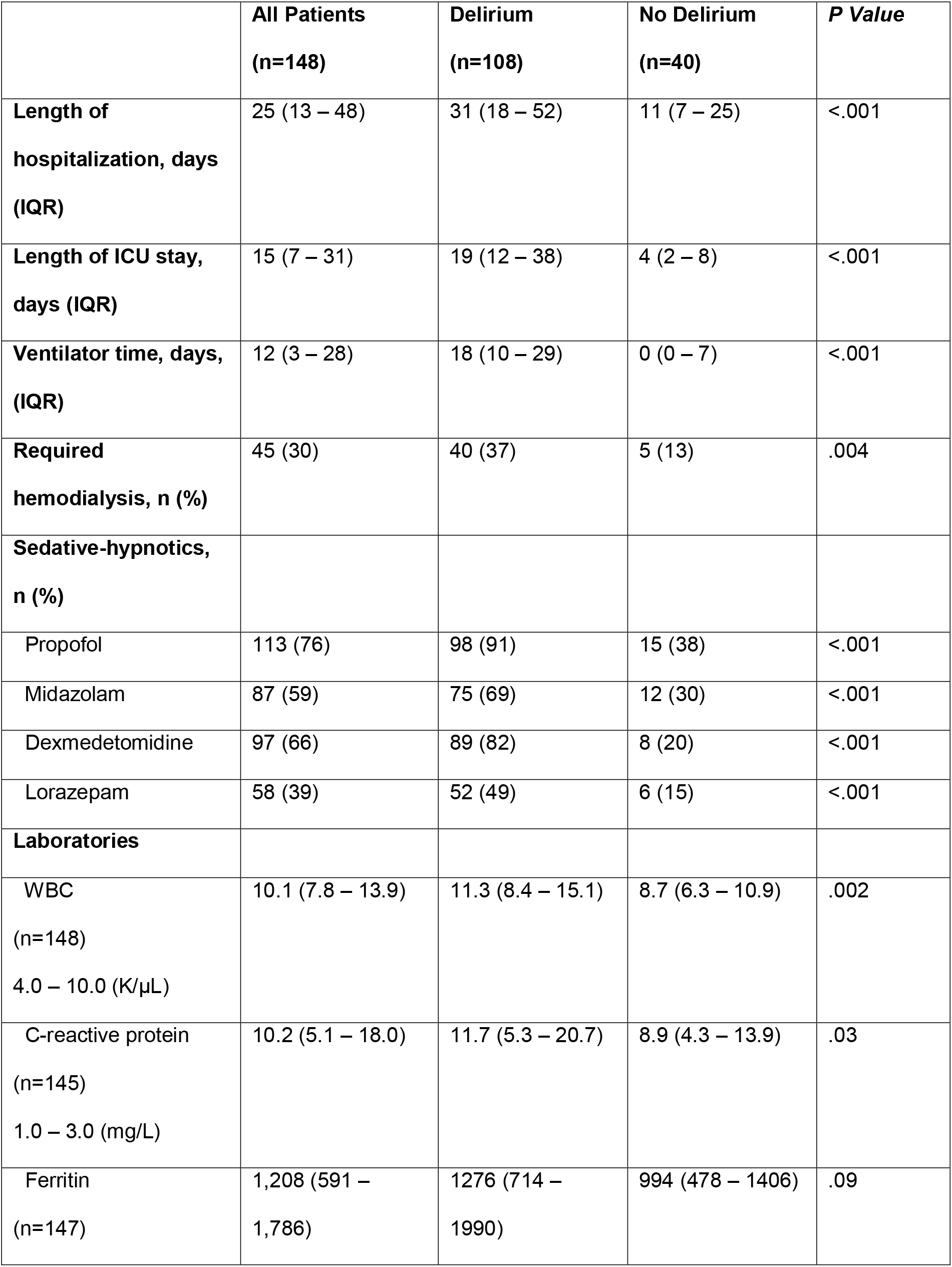
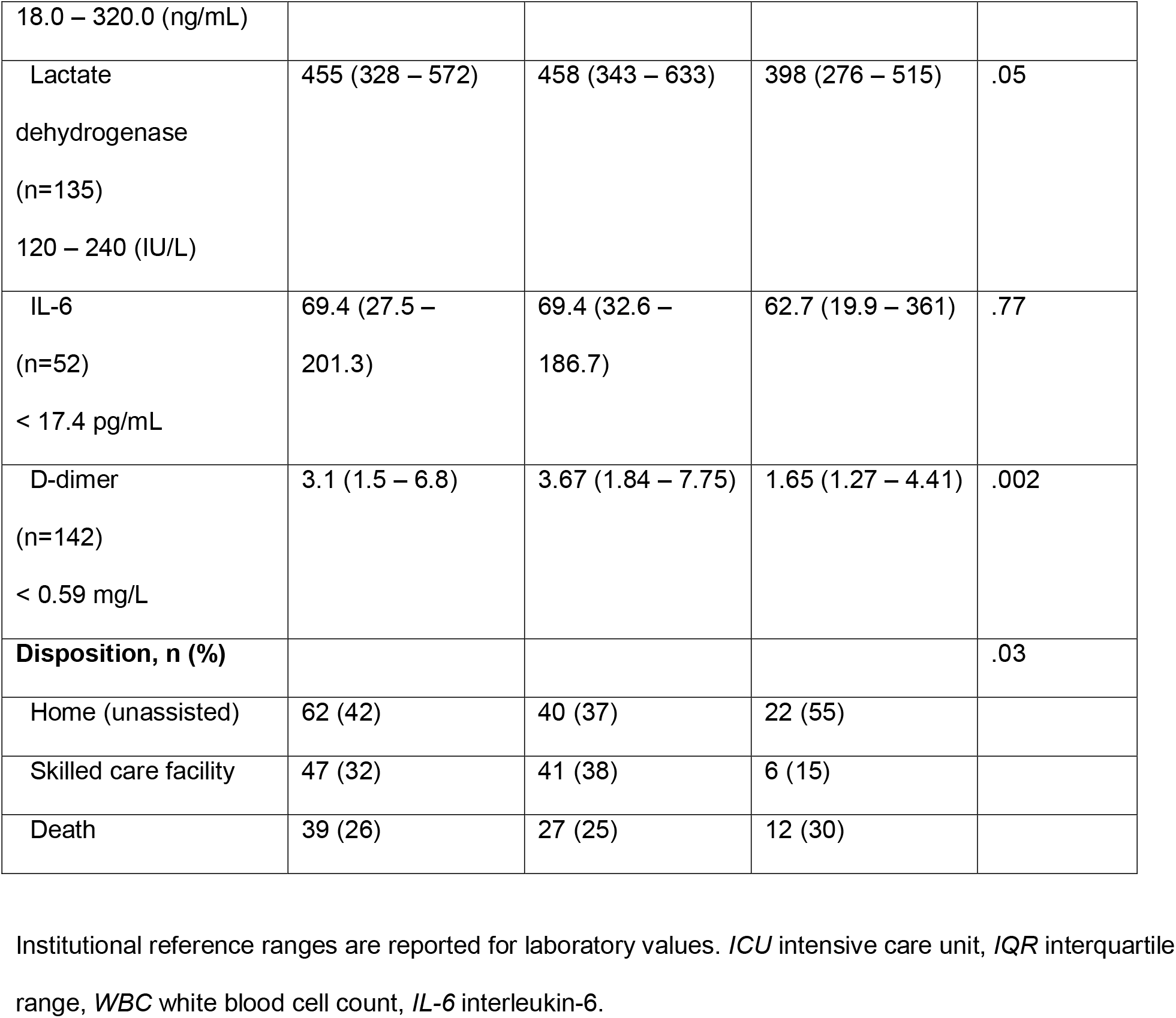
Hospitalization

|  | **All Patients**<br>**(n=148)** | **Delirium**<br>**(n=108)** | **No Delirium**<br>**(n=40)** | **P Value** |
| --- | --- | --- | --- | --- |
| **Length of hospitalization, days (IQR)** | 25 (13 – 48) | 31 (18 – 52) | 11 (7 – 25) | <.001 |
| **Length of ICU stay, days (IQR)** | 15 (7 – 31) | 19 (12 – 38) | 4 (2 – 8) | <.001 |
| **Ventilator time, days, (IQR)** | 12 (3 – 28) | 18 (10 – 29) | 0 (0 – 7) | <.001 |
| **Required hemodialysis, n (%)** | 45 (30) | 40 (37) | 5 (13) | .004 |
| **Sedative-hypnotics, n (%)** |  |  |  |  |
| Propofol | 113 (76) | 98 (91) | 15 (38) | <.001 |
| Midazolam | 87 (59) | 75 (69) | 12 (30) | <.001 |
| Dexmedetomidine | 97 (66) | 89 (82) | 8 (20) | <.001 |
| Lorazepam | 58 (39) | 52 (49) | 6 (15) | <.001 |
| **Laboratories** |  |  |  |  |
| WBC<br>(n=148)<br>4.0 – 10.0 (K/μL) | 10.1 (7.8 – 13.9) | 11.3 (8.4 – 15.1) | 8.7 (6.3 – 10.9) | .002 |
| C-reactive protein<br>(n=145)<br>1.0 – 3.0 (mg/L) | 10.2 (5.1 – 18.0) | 11.7 (5.3 – 20.7) | 8.9 (4.3 – 13.9) | .03 |
| Ferritin<br>(n=147) | 1,208 (591 – 1,786) | 1276 (714 – 1990) | 994 (478 – 1406) | .09 |
| 18.0 – 320.0 (ng/mL) |  |  |  |  |
| Lactate<br>dehydrogenase<br>(n=135)<br>120 – 240 (IU/L) | 455 (328 – 572) | 458 (343 – 633) | 398 (276 – 515) | .05 |
| IL-6<br>(n=52)<br>< 17.4 pg/mL | 69.4 (27.5 –<br>201.3) | 69.4 (32.6 –<br>186.7) | 62.7 (19.9 – 361) | .77 |
| D-dimer<br>(n=142)<br>< 0.59 mg/L | 3.1 (1.5 – 6.8) | 3.67 (1.84 – 7.75) | 1.65 (1.27 – 4.41) | .002 |
| **Disposition, n (%)** |  |  |  | .03 |
| Home (unassisted) | 62 (42) | 40 (37) | 22 (55) |  |
| Skilled care facility | 47 (32) | 41 (38) | 6 (15) |  |
| Death | 39 (26) | 27 (25) | 12 (30) |  |

Institutional reference ranges are reported for laboratory values. *ICU* intensive care unit, *IQR* interquartile range, *WBC* white blood cell count, *IL-6* interleukin-6.

Neuropsychological outcomes after discharge are reported in Table 4. Among patients who were still alive and available to complete survey materials, nearly 25% of patients (4/17) scored positive for delirium based on family assessment (FAM-CAM), and all of these patients were delirious during hospitalization. Similarly, approximately 23% of patients (5/22) demonstrated either questionable impairment or impairment consistent with dementia based on the Short Blessed Test, and all five of these patients were also delirious during hospitalization. Of note, three of these five patients also screened positive for delirium based on the FAM-CAM. Lastly, 12% of patients (3/25) screened positive for depression after discharge. The three patients who screened positive also experienced delirium during ICU admission.

**Table 4.** Post-Discharge Neuropsychological Outcomes^**†**^

|  | **All Patients**<br>**(n=148)** | **Delirium**<br>**(n=108)** | **No Delirium**<br>**(n=40)** |
| --- | --- | --- | --- |
| Positive FAM-CAM, n (%)<br>(n=17) | 4 (24) | 4 (31) | 0 (0) |
| PROMIS 4A Cognitive<br>Abilities Score, median<br>(IQR)<br>(n=25) | 16 (10 – 20) | 17 (9 – 20) | 14 (6) |
| Short Blessed Test –<br>Normal, n (%)<br>(n=22) | 17 (74) | 10 (67) | 7 (100) |
| Short Blessed Test –<br>questionable cognitive<br>impairment, n (%)<br>(n=22) | 2 (8.7) | 2 (13) | 0 (0) |
| Short Blessed Test –<br>cognitive impairment, n (%)<br>(n=22) | 3 (13) | 3 (20) | 0 (0) |
| PHQ-9 screen positive, n,<br>(%)<br>(n=25) | 3 (12) | 3 (17) | 0 (0) |

*FAM-CAM* Family-based Confusion Assessment Method for delirium, *PROMIS* Patient-Reported Outcomes Measurement Information System, *PHQ* Patient Health Questionnaire. †For each post-discharge survey, proportions are calculated based on the total numbers of surveys completed. In total, 25 surveys were completed for the PROMIS 4A test, 25 surveys were completed for the PHQ-9, 17 surveys were completed for the FAM-CAM, and 22 surveys were completed for the Short Blessed Test.
Reasons for not completing a test included the following: unable to contact (n=54), patient deceased (n=43), refusal (n=18), unable to provide consent (n=5), and admission to skilled care facility (n=3).

### Neuroradiological Findings

In total, 47 patients underwent neuroimaging during hospitalization. The majority of imaging results were unremarkable or demonstrated incidental findings unrelated to COVID-19. However, some notable findings were present. A brain MRI was ordered for a 59-year-old female with COVID-19 pneumonia and worsening encephalopathy (i.e., no response to commands or noxious stimulus). Imaging revealed abnormal fluid attenuated inversion recovery (FLAIR) hyperintensity affecting the occipital and temporal lobes (Figures 1A, 1B), microhemorrhage in the splenium of the corpus callosum (Figures 1B, 1C) and posterior leptomeningeal enhancement Figures 1C, 1D), suggestive of encephalitis. A 29-year-old female with history of ovarian malignancy presented with new-onset seizures, for which an MRI was ordered. Results revealed diffuse dural thickening and enhancement (Supplemental Figure 1A) one day prior to positive COVID testing. The differential diagnosis included intracranial hypotension (recent lumbar puncture), inflammation, infection, and neoplastic processes. No definitive diagnosis was reached, though this enhancement resolved approximately one month later (Supplemental Figure 1B). Lastly, one patient demonstrated diffuse parenchymal abnormalities on MRI suggestive of bilateral hypoxic-ischemic injury (Supplemental Figure 2). This patient experienced two separate arrests (pulseless electrical activity) within the preceding three weeks. A non-contrast head CT two weeks later demonstrated poor sulcation bilaterally, suggesting global hypoxic-ischemic injury (Supplemental Figure 3).

**Figure 1.**
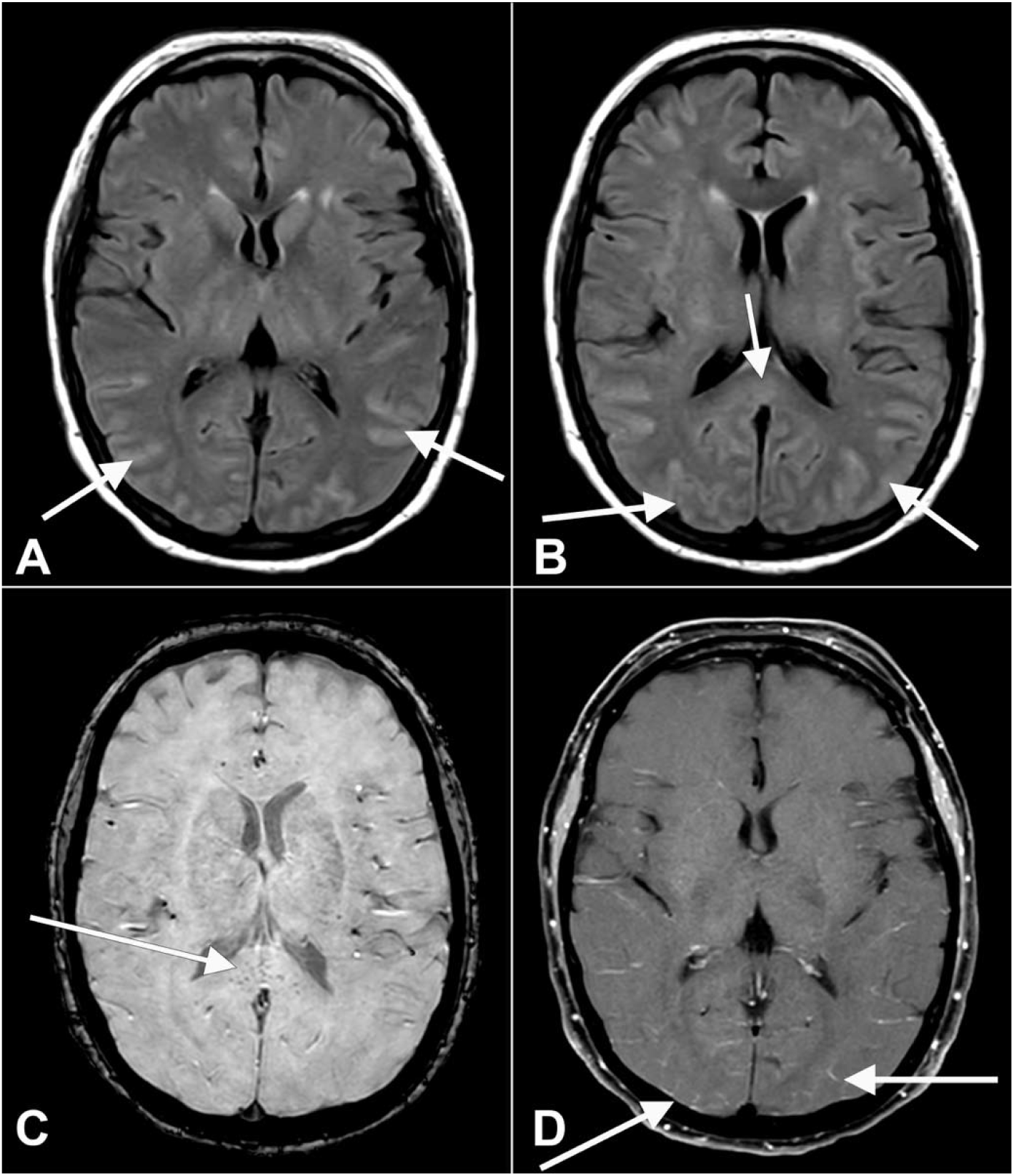
Axial fluid-attenuated inversion recovery (FLAIR) (A, B) images at the level of the basal ganglia show abnormal FLAIR hyperintense signal (arrows) affecting the bilateral occipital, temporal lobes. This appears almost sulcal suggesting a higher protein component within the cerebrospinal fluid. Note the elevated FLAIR signal in the splenium of the corpus callosum (arrow) suggesting parenchymal insult. Axial susceptibility weighted imaging (SWI) (C) at the level of the splenium of the corpus callosum shows small areas of susceptibility (arrow) in the splenium, likely related to microhemorrhage. Axial T1 (D) post-contrast with fat suppression at the level of the basal ganglia shows subtle, though true, enhancement (arrows) in the posterior sulci, arachnoid pial (leptomeningeal) pattern suggesting a degree of encephalitis.

## Discussion

In a cohort of ICU patients with COVID-19, delirium was a common complication, affecting more than 70% of patients. Furthermore, delirium was associated with prolonged hospitalization, increased length of ICU stay, discharge to skilled care facilities, and positive screens for neuropsychological impairment up to two months after discharge. Delirium occurred in the setting of multiple sedative-hypnotic agents, acute inflammatory responses, deviation from delirium prevention protocols, and cerebrovascular events, which are all factors that could have further catalyzed delirium precipitation. ICU liberation activities were infrequently implemented compared to the protocolized frequency expected. Overall, the burden of cognitive impairment was high in patients with COVID-19, as was the risk of related complications.

These results align with previous data demonstrating a high incidence of delirium in critically ill patients with COVID-19.^1-4^ Our findings also highlight the multifactorial nature of delirium risk factors. In terms of demographics, 50% of patients in the delirium group were African American. COVID-19 has adversely, and disproportionately, impacted racial and ethnic minority communities,^20, 21^ and our results further suggest an increased risk of attendant complications (e.g., delirium) during hospitalization. Efforts to reduce racial healthcare disparities may thus, by extension, mitigate risk of delirium and related consequences of COVID-19. Patients experiencing delirium also demonstrated significantly increased weight, and obesity may drive organ dysfunction via immune system dysregulation.^22^ Additionally, there was a disproportionate number of female patients in the delirium group (absolute standardized difference >.30). These results are discrepant from a prior case series of critically ill patients with COVID-19 demonstrating an increased risk of delirium with male patients.^4^ Male sex has also been identified as an independent risk factor for delirium in other patient populations, possibly due to underlying comorbidity severity.^23, 24^ Whether the findings in this study are spurious or reflect an underlying biological phenomenon is unclear. Further investigation is warranted to improve understanding of the impact that such demographic factors on delirium risk in patients with COVID-19.

Cognitive dysfunction may also occur as a result of direct coronavirus invasion of the central nervous system^6^ or other indirect mechanisms, such as polypharmacy, systemic inflammatory responses, and cerebrovascular events. Indeed, benzodiazepine sedation was common in this patient cohort, with nearly 60% of patients receiving midazolam at one point during ICU admission. Lorazepam was a common sedation agent as well, and benzodiazepine use is associated with delirium in critically ill patients.^25-27^ Whether benzodiazepine administration served as a driver of delirium, or reflected worsening agitation (prompting additional sedative agents), remains unclear. Inflammation may have also contributed to delirium risk. Inflammatory markers (e.g., c-reactive protein, ferritin, interleukin-6, lactate dehydrogenase) were considerably elevated in this patient cohort. In fact, serum levels observed in this study aligned with – or exceeded – previously reported values in patients with severe COVID-19,^2, 5^ and there was MRI evidence of neuroinflammation for at least two patients in this series. C-reactive protein was elevated in delirious patients, and c-reactive protein increases blood-brain barrier permeability in basic science models.^28^ However, this was an unadjusted, bivariable analysis, and confounding remains possible. Cerebral ischemia may also contribute to delirium in patients with COVID-19. Severe hypoxic-ischemic injury occurred in a patient who experienced multiple cardiopulmonary arrests during the course of illness. Stroke has previously been reported in patients with COVID-19,^29^ as thromboembolic phenomena and cerebral malperfusion may both occur during the clinical course of COVID-19. Lastly, overall illness severity may increase delirium risk. Indeed, patients with delirium had prolonged hospital and ICU lengths of stay, longer duration of mechanical ventilation, and were more likely to require hemodialysis. Overall, multiple processes likely contribute to delirium in patients with COVID-19. Targeted case-control studies can be conducted to determine independent risk factors for delirium in this patient population.

Delirium prevention and management are inherently challenging for COVID-19 patients. While delirium prevention bundles have been demonstrated to reduce risk,^30, 31^ unique challenges posed by COVID-19 hinder the implementation of standard prevention practices. Spontaneous awakening and breathing trials, for example, may not have been possible due to illness severity and associated ventilator requirements. Clinicians may have also been limited in terms of sedation regimen. Agitation was commonly observed, and nearly 30% of patients required antipsychotics in this cohort. Agitation and hyperactive delirium likely prompted additional sedation and prolonged use of physical restraints. Early mobility was limited given illness severity, and family engagement was not possible due to visitor policy restrictions. In-person interactions with clinicians were also limited given the intent of reducing virus transmission. As such, the culmination of disease severity, limited face-to-face time spent with patients, and visitor restriction policies likely hindered ICU liberation bundle implementation. Novel strategies for implementing delirium prevention bundles in this patient population may help to further mitigate risk and should be tested in prospective trials.

Neuropsychological impairment after discharge was also present for some patients based on subjective reporting, caretaker assessment, and objective testing for depression and cognitive impairment. Furthermore, all patients that screened positive for possible impairment also experienced delirium in the hospital. These estimates may have been even higher, given that many patients called for post-discharge assessments were unable to be reached, refused participant, or were still admitted to skilled care facilities. Whether post-discharge cognitive impairment was related specifically to COVID-19 or critical illness more broadly is unclear. Indeed, cognitive impairment is common at discharge for patients who experienced delirium while in the ICU, and delirium is present for nearly 20% of patients newly admitted to acute care facilities.^7, 32^ Moreover, cognitive impairment can be present for months-to-years after acute respiratory distress syndrome and sepsis,^33-35^ and symptoms of depression and post-traumatic stress disorder are commonly reported among ICU survivors.^36^ Neuropsychological impairment after discharge may thus, in part, reflect critical illness, rather than pathophysiologic insults specific to COVID-19. Nonetheless, ICU patients with COVID-19 experience considerable neuropsychological burden both during and after hospitalization. Identifying such vulnerable patients will be important for providing appropriate longitudinal care and resources.

The strengths of this study include granular data with respect to delirium, identification of potential risk factors, characterization of delirium prevention strategies, and post-discharge outcomes. Data were representative of an academic tertiary care center with nearly 150 patients. A validated, standardized chart review method was used to identify delirium,^18^ and the study measures used to characterize cognitive function, such as the FAM-CAM, Short Blessed Test, and PROMIS assessments, are standard measures that increase confidence in the results. In terms of limitations, this is a this was a single center analysis, and the results are restricted to the institution studied. The study was not conducted with a matched control cohort, as this was a descriptive study. The post-discharge telephone-based assessments served as screening tools rather than thorough neuropsychological testing batteries. As such, these post-discharge results are preliminary and warrant rigorous, follow-up analysis. Additionally, neuropsychological impairment may have been present at baseline for some patients, particularly for those with previous neurological disorders. Baseline neuropsychological testing was not possible for this study. Lastly, data were limited for post-discharge cognitive outcomes, as more than half of patients called were unavailable to complete assessments.

In summary, delirium is common complication of COVID-19 with multiple contributing factors. Furthermore, neuropsychological impairment may persist in some patients after discharge. Further research should aim to identify independent risk factors in this population and novel, effective prevention strategies.

## Data Availability

All de-identified data will be made available upon request.

## Abbreviations and Acronyms

ABCDEF: Assess, prevent and manage pain; both spontaneous awakening and breathing trials; choice of analgesia and sedation; delirium: assess, prevent, and manage; early mobility and exercise; family engagement and empowerment
COVID-19: coronavirus disease 2019
FAM-CAM: family confusion assessment method
FLAIR: fluid attenuated inversion recovery
ICU: intensive care unit
MRI: magnetic resonance imaging
PROMIS: patient-reported outcomes measurement information system
SARS-CoV-2: severe acute respiratory syndrome coronavirus 2
SWI: susceptibility weighted imaging

## Acknowledgments

We would like to acknowledge Dr. Michael Kenes (PharmD, BCPS, BCCCP) and Ms. Margaret Diehl for assistance with medical chart data extraction.

