## Supplemental Online Material for "Delirium and Neuropsychological Outcomes in Critically Ill Patients with COVID-19: an Institutional Case Series"

CARE case report guidelines

### CARE Checklist of information to include when writing a case report

(cc) BY-NC-ND

| Topic | Item | Checklist item description | Reported on Line |
|---|---|---|---|
| Title | 1 | The diagnosis or intervention of primary focus followed by the words "case report" | Pg 1., Lines 1-2 (case series) |
| Key Words | 2 | 2 to 5 key words that identify diagnoses or interventions in this case report, including "case report" | Pg 4, Lines 60-61 |
| Abstract (no references) | 3a | Introduction: What is unique about this case and what does it add to the scientific literature? | Pgs. 3-4 |
| | 3b | Main symptoms and/or important clinical findings | Pg. 3, Lines 43-53 |
| | 3c | The main diagnoses, therapeutic interventions, and outcomes | Pg. 3, Lines 43-53 |
| | 3d | Conclusion—What is the main "take-away" lesson(s) from this case? | Pg. 4, Lines 55-57 |
| Introduction | 4 | One or two paragraphs summarizing why this case is unique (**may include references**) | Pgs. 6-7 |
| Patient Information | 5a | De-identified patient specific information. | Pgs. 12-14, Tables 1-4, Figures |
| | 5b | Primary concerns and symptoms of the patient. | Pg. 12, Lines 196-209 |
| | 5c | Medical, family, and psycho-social history including relevant genetic information | Table 1 |
| | 5d | Relevant past interventions with outcomes | Table 2 |
| Clinical Findings | 6 | Describe significant physical examination (PE) and important clinical findings. | Tables 2-4 |
| Timeline | 7 | Historical and current information from this episode of care organized as a timeline | N/A |
| Diagnostic Assessment | 8a | Diagnostic testing (such as PE, laboratory testing, imaging, surveys). | Pgs 9-10, 14 |
| | 8b | Diagnostic challenges (such as access to testing, financial, or cultural) | N/A |
| | 8c | Diagnosis (including other diagnoses considered) | Table 2, pg. 12, line 197 |
| | 8d | Prognosis (such as staging in oncology) where applicable | Pg. 13, lines 222-231 |
| Therapeutic Intervention | 9a | Types of therapeutic intervention (such as pharmacologic, surgical, preventive, self-care) | Table 2, pg. 12, lines 201-208 |
| | 9b | Administration of therapeutic intervention (such as dosage, strength, duration) | Table 2, pg. 12, lines 201-208 |
| | 9c | Changes in therapeutic intervention (with rationale) | Table 2, pg. 12, lines 201-208 |
| Follow-up and Outcomes | 10a | Clinician and patient-assessed outcomes (if available) | Table 4 (patient/caregiver assessment) |
| | 10b | Important follow-up diagnostic and other test results | Table 4 |
| | 10c | Intervention adherence and tolerability (How was this assessed?) | Table 2, pg. 12, lines 201-205 |
| | 10d | Adverse and unanticipated events | Tables 2-4 (outcomes, disposition) |
| Discussion | 11a | A scientific discussion of the strengths AND limitations associated with this case report | Pg. 19, lines 347-363 |
| | 11b | Discussion of the relevant medical literature **with references**. | Pg. 15-18 |
| | 11c | The scientific rationale for any conclusions (including assessment of possible causes) | Pg. 17, lines 307-310 |
| | 11d | The primary "take-away" lessons of this case report (without references) in a one paragraph conclusion | Pg. 13=9, lines 365-368 |
| Patient Perspective | 12 | The patient should share their perspective in one to two paragraphs on the treatment(s) they received | Table 4 (survey outcomes) |
| Informed Consent | 13 | Did the patient give informed consent? Please provide if requested | Yes ☒* No ☐ |

*Yes, pertaining to post-discharge telephone surveys.

**Supplemental Figure 1**

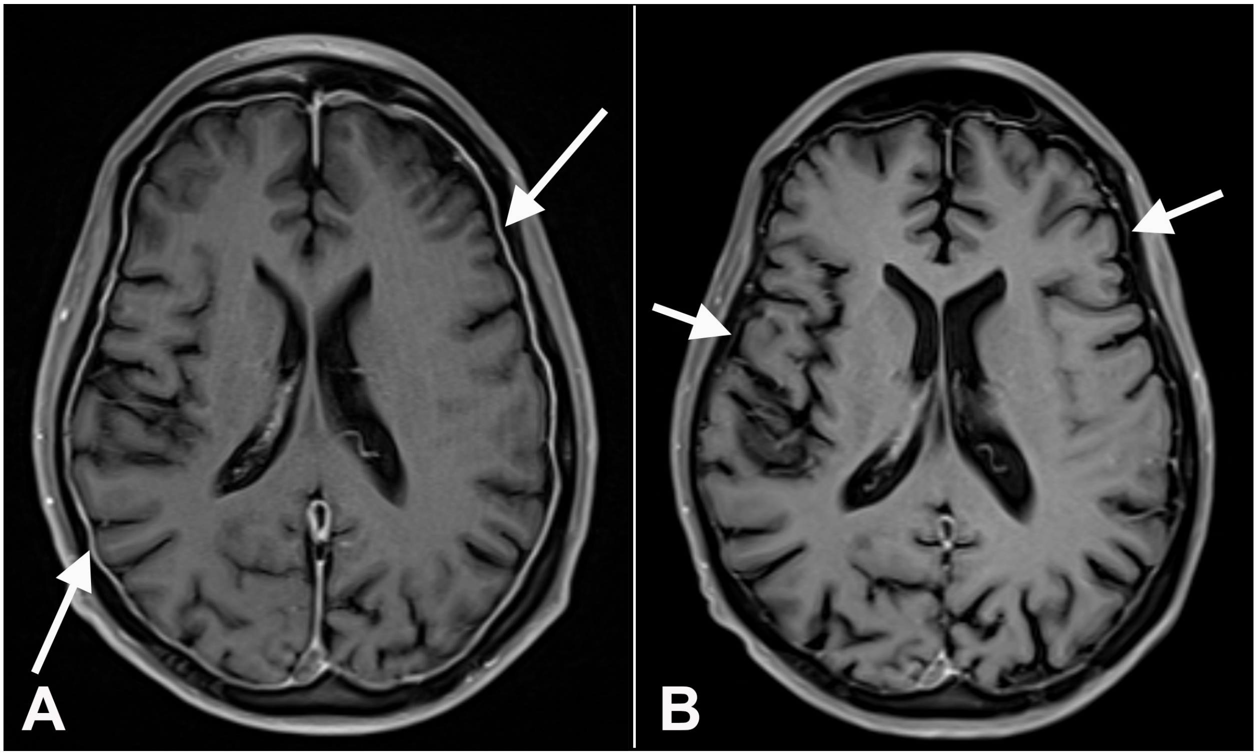


A, Axial T1 post-contrast with fat suppression images at the level of the mid lateral ventricles demonstrate smooth dural enhancement (arrows) along the bilateral cerebral convexities. B, This feature resolves one month later (arrows). The overall pattern of dura-arachnoid/ pachymeningeal enhancement is non-specific, can be seen with intracranial hypotension, in the procedural setting (e.g., lumbar puncture), and other scenarios (e.g., infection, inflammation).

**Supplemental Figure 2**

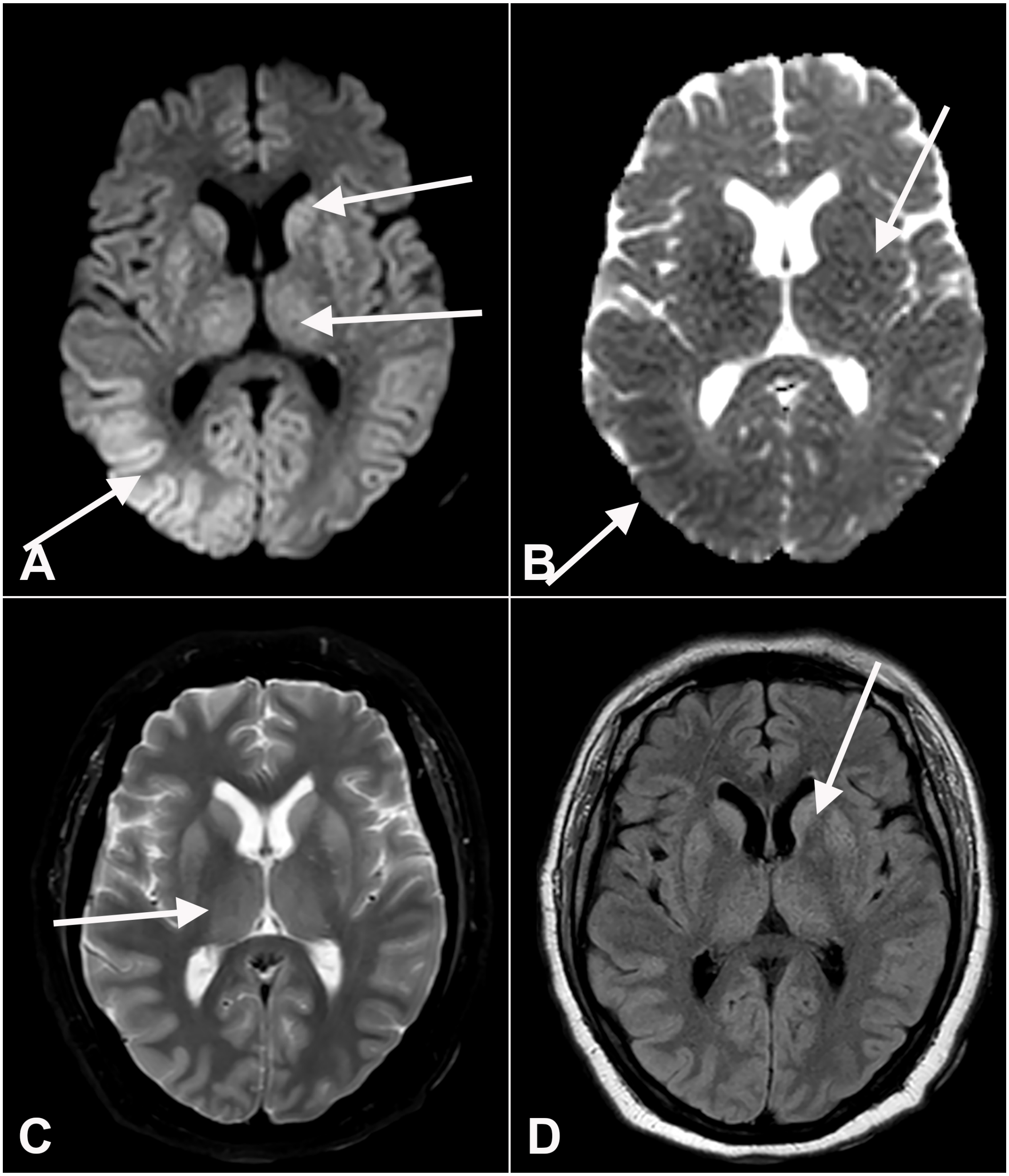


After initial non-contrast head CT (which was unremarkable), the patient had diffuse parenchymal abnormalities on MRI examination. A, Axial diffusion-weighted imaging shows hyperintense signal (arrows) at the bilateral basal ganglia, thalami, and posterior cortices, regions are hypointense (arrows) on corresponding apparent diffusion coefficient map (B). These same locations are hyperintense on T2 (C) and FLAIR (D), especially at the basal ganglia and thalami.

#### Supplemental Figure 3

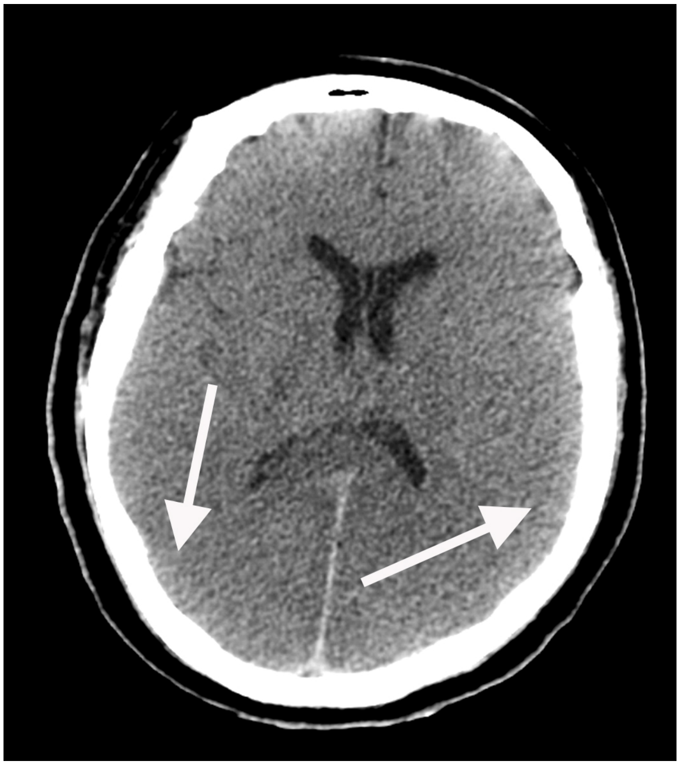

Non-contrast head CT performed approximately two weeks after cardiopulmonary arrest shows poor sulcation (arrows) bilaterally, suggesting global insult, most likely hypoxic-ischemic in etiology
